# Outcomes of Antiplatelet Therapy Continuation in Older Hypertensive Adults With Peptic Ulcer Disease

**DOI:** 10.1101/2023.12.11.23299836

**Authors:** Yi-Tsang Lan, Kar-Chun Lim, Chung-Yu Ho, Ying-Ting Chao, Tsung-Yi Yen, Ming-Feng Shih, Chien-Hsieh Chiang

## Abstract

**Background:** The appropriateness of continuation of antiplatelet therapy in older hypertensive aspirin users with documented peptic ulcer disease (PUD) is uncertain.

**Methods:** This multicenter cohort study screened adults aged 65 years or older, using aspirin for primary and secondary cardiovascular disease prevention between January 2014 and December 2018. Patients with panendoscopy-proven PUD and hypertension were identified. Subsequent antiplatelet strategies were categorized as aspirin discontinuation (AD), aspirin continuation (AC), and switch to clopidogrel (SC) groups. Inverse probability of treatment weighting was applied to balance baseline characteristics. The main outcomes were incident major adverse cardiac events (MACEs) and hospitalizations for upper gastrointestinal bleeding (UGIB), followed through 31 December 2020.

**Results:** 735 eligible patients were analyzed. During a median follow-up of 39.7 months, 178 MACEs occurred. Compared with AD, SC was not related to the risk of incident MACEs, but AC increased the risk of incident MACEs (adjusted HR, 1.58; 95% CI, 1.04-2.38) in secondary prevention patients. On the other hand, 102 hospitalizations for UGIB occurred during a median follow-up of 43.4 months. Compared with AD, neither AC nor SC affected the risk of hospitalization for UGIB in secondary prevention patients. However, secondary prevention patients with chronic kidney disease were at increased risk of hospitalizations for UGIB (adjusted HR, 2.41; 95% CI, 1.30-4.47).

**Conclusions:** AC may increase the risk of incident MACEs in older hypertensive adults with PUD previously taking aspirin for secondary cardiovascular disease prevention. The appropriateness of antiplatelet therapy continuation after PUD is diagnosed in older hypertensive adults warrants rigorous considerations.

## Introduction

Aspirin is recommended for secondary prevention of cardiovascular disease (CVD), including acute coronary syndrome, ischemic stroke, unstable or stable angina, and peripheral arterial disease.^1–3^ However, it is controversial if aspirin should be used for the primary prevention of CVD. Although age is an independent risk factor for atherosclerotic CVD,^4^ large trials failed to reveal protective effects of aspirin for primary prevention of atherosclerotic CVD in older adults, even in patients with cardiovascular risk factors.^5–7^ Hypertension is one of the most common cardiovascular risk factors, and its prevalence also increases with age, estimated to affect 74.5% of the older population in the United States (US) and 63% in Taiwan.^8,9^ Aspirin is widely used in older hypertensive patients for CVD protection because both hypertension and aging lead to CVDs. Nonetheless, the appropriateness of aspirin treatment in older hypertensive individuals should be adjudicated by considering both the effectiveness of CVD prevention and the risk of bleeding events.

Long-term aspirin use in older hypertensive people may raise the concern of an increased risk of hemorrhagic events, such as gastrointestinal bleeding and intracranial hemorrhage.^10,7,11^ An early study demonstrated bleeding risks of peptic ulcer disease (PUD) were raised for all doses of aspirin taken (75-300 mg daily).^12^ Other reports showed that both low-dose aspirin and enteric-coated aspirin increased the risk of upper gastrointestinal bleeding (UGIB).^13,14^ In community-dwelling older adults without CVD, daily aspirin was observed to increase the rate of major hemorrhage without benefits to disability-free survival or CVD prevention.^7,15^ Discontinuation of aspirin therapy in patients with UGIB is a reasonable “do-no-harm” practice, but long-term aspirin use is still suggested in patients taking aspirin for secondary CVD prevention to reduce mortality.^16,17^ A randomized clinical trial of low-dose aspirin users with UGIB concluded that continuous aspirin therapy reduced the mortality rate within 8 weeks after endoscopic control of peptic ulcer bleeding, but increased the risk of recurrent bleeding within 30 days.^17^ In addition, both age and hypertension were shown to increase the risk of UGIB in older adults taking aspirin for secondary CVD prevention.^18,19^ However, no large-sized clinical trials have examined if older hypertensive adults with documented peptic ulcer disease (PUD) should keep taking aspirin for CVD prevention.

This study aimed to investigate the outcomes of antiplatelet drug use in older hypertensive patients taking aspirin for primary or secondary CVD prevention who develop PUD. The outcomes of interest were major adverse cardiovascular events (MACEs) and hospitalizations for UGIB.

## Methods Study

### Design

This retrospective multicenter cohort study was conducted at the National Taiwan University Hospital (NTUH) and its branches in Northern, Central, and Southern Taiwan. The medical records were accessed from the NTUH-Integrative Medical Database (**Supplemental Methods**). The study protocol was approved by the NTUH Research Ethics Committee (201907023RINA), and the requirement of informed patient consent was waived. This study was performed in accordance with the STROBE guidelines.

### Patients

The records of all aspirin users aged 65 years or older who received a panendoscopy between 1 January 2014 and 31 December 2018 at the NTUH Healthcare System were accessed (**Supplemental Figure S1**). We reviewed all panendoscopy reports of 4550 patients to identify patients with confirmed PUD. The diagnosis date of panendoscopy-proven PUD served as the index date. Excluded were non-hypertensive patients, patients with concomitant use of warfarin or novel oral anticoagulants, patients without documented PUD, and dual antiplatelet users before or after index dates.

All eligible hypertensive patients were classified according to the use of aspirin for primary and secondary CVD prevention. The baseline groups were categorized according to the administration of antiplatelet drugs within 3 months after a panendoscopy diagnosis of PUD. Aspirin discontinuation (AD) was defined as taking aspirin for < 28 days within 3 months after the index date. Aspirin continuation (AC) was defined as taking aspirin for ≥ 28 days within 3 months after the index date. In the AC group, the total amount of aspirin taken within 12 months after the index date based was calculated based on the daily aspirin dose. Switch to clopidogrel (SC) was defined as taking clopidogrel for ≥ 28 days. The single-payer National Health Insurance Program in Taiwan reimburses for the cost of clopidogrel for secondary prevention of CVD in patients with panendoscopy-proven PUD or aspirin allergy.

Patient demographic data extracted from the medical records included age and sex. Comorbidities were identified by ICD-10 codes (**Supplemental Methods**), including chronic kidney disease (CKD), diabetes, liver cirrhosis, heart failure, and Helicobacter *pylori* (HP) infection. The score of the Charlson comorbidity index (CCI) was calculated. Concomitant medications included non-steroidal anti-inflammatory drugs, antihypertensive drugs, lipid-lowering agents, oral antidiabetic drugs, insulin analogs, and proton pump inhibitors (PPIs) prescribed for 28 days or more within 6 months before the index date. PPI use after a diagnosis of PUD was also examined.

### Main Outcome Measures

The main outcomes were incident MACEs and hospitalizations for UGIB after the index date. MACEs included cardiovascular death, non-fatal myocardial infarction, and non-fatal stroke after the index date. Hospitalizations for UGIB were confirmed by documentation of endoscopic reports and diagnosis codes during hospitalizations after the index date. Death registry files were obtained from the government’s annual statistics data. Outcomes were examined until 31 December 2020.

### Statistical Analysis

Categorical data were compared by the chi-squared test or Fisher’s exact test. Continuous data were compared using the independent t-test. Kaplan–Meier failure plots of the 3 baseline groups were drawn. Inverse probability of treatment weighting (IPTW) was applied to balance baseline group characteristics by a weight of propensity scores.^20^ Propensity scores were estimated by using a logistic regression model of all covariates (**Supplemental Methods**). Cox proportional hazards regression analysis of the IPTW population was performed to determine the association of subsequent antiplatelet drug use with incident MACEs and hospitalizations for UGIB. Models contained subsequent antiplatelet drugs, age, sex, CCI, comorbidities, HP infection, and concomitant medications. The proportionality assumption was tested and met. We assumed missing values over time were missing at random and performed listwise deletion. All statistical analyses were 2-tailed, and a value of *P* < .05 was considered to indicate statistical significance. Statistical analyses were performed with SAS version 9.4 software (SAS Institute Inc., Cary, NC, USA).

## Results

### Patient Characteristics

Among 735 older hypertensive patients included in the analysis, 346 (47.1%) were male, the mean age was 77.3 ± 7.5 years, 378 (51.4%) patients used aspirin for primary CVD prevention, and 357 (48.6%) patients used aspirin for secondary CVD prevention (**Figure S1**). After IPTW, the patients previously on aspirin for primary CVD prevention had a more balanced distribution of age, sex, CCI, type 2 diabetes, malignancy, heart failure, and PPI use between AD and AC groups (**Table S1**); the patients previously prescribed aspirin for secondary CVD prevention had a more balanced distribution of age, sex, CCI, type 2 diabetes, liver cirrhosis, and PPI use between AD and AC groups (**Table S2**).

### Incident MACEs

During a median follow-up period of 39.7 months, 178 incident MACEs occurred in the IPTW population. Subsequent antiplatelet strategies in patients previously on aspirin for primary CVD prevention were not associated with a cumulative risk of incident MACEs in Kaplan–Meier failure plots whether before IPTW (*P* = .301) (**Figure S2A**) or after IPTW (*P* = .251) (**Figure 1A**). In the multivariate Cox proportional hazards model of the IPTW population, neither AC (adjusted HR, 1.58; 95% CI, 0.75-3.34) nor SC (adjusted HR, 1.29; 95% CI, 0.33-4.98) was associated with a reduced risk of incident MACEs in patients previously prescribed aspirin for primary CVD prevention, compared with AD (**Table 1**). On the other hand, subsequent antiplatelet strategies in patients previously on aspirin for secondary CVD prevention were not related to the cumulative risk of incident MACEs in Kaplan–Meier failure plots whether before IPTW (*P* = .180) (**Figure S3A**) or after IPTW (*P* = .170) (**Figure 2A**). While SC was still not related to a lowered risk of incident MACEs, AC increased the risk of incident MACEs in patients previously prescribed aspirin for secondary CVD prevention (adjusted HR, 1.58; 95% CI, 1.04-2.38), compared with AD in the multivariate Cox proportional hazards analysis of the IPTW population (**Table 2**).

**Figure 1.**
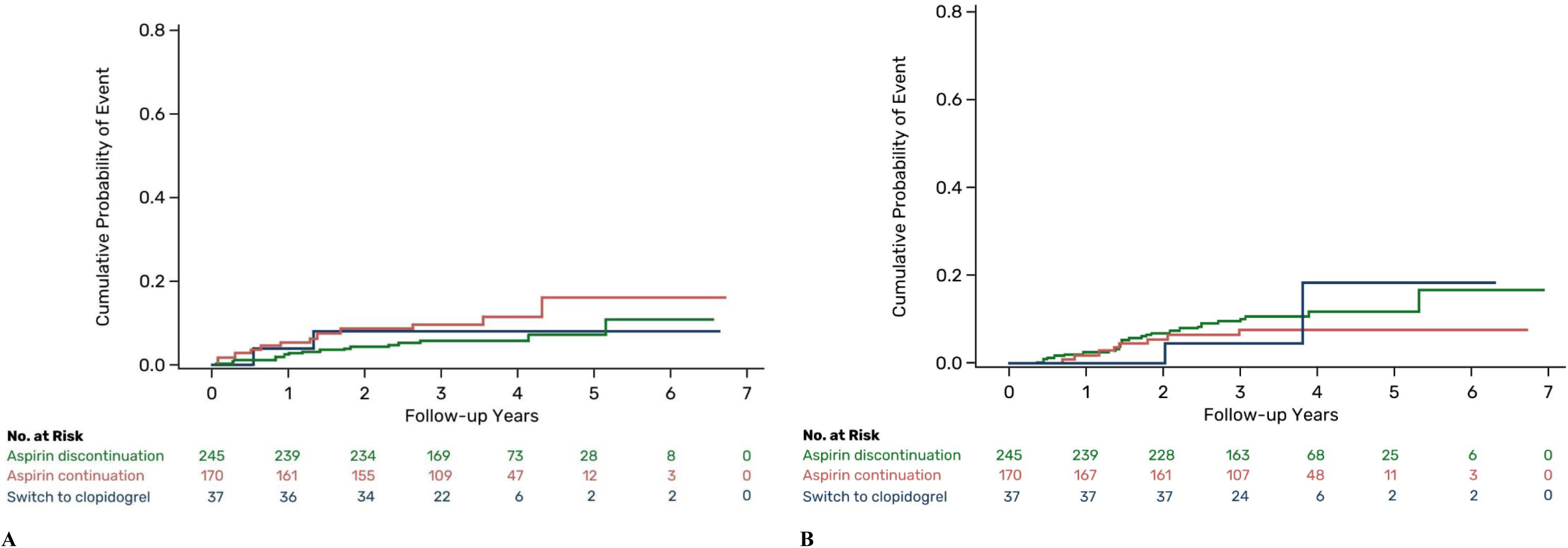
Association of subsequent antiplatelet strategies with MACEs and hospitalizations for UGIB in patients previously on aspirin for primary CVD prevention. Kaplan–Meier failure plots show cumulative risks of major adverse cardiovascular events (**A**) and hospitalizations for upper gastrointestinal bleeding (**B**) of the three groups after inverse probability treatment weighting. The log-rank tests were not significant with *P* = .251 (**A**) and *P* = .630 (**B**). CVD, cardiovascular disease; MACE, major adverse cardiac event; UGIB, upper gastrointestinal bleeding.

**Figure 2.**
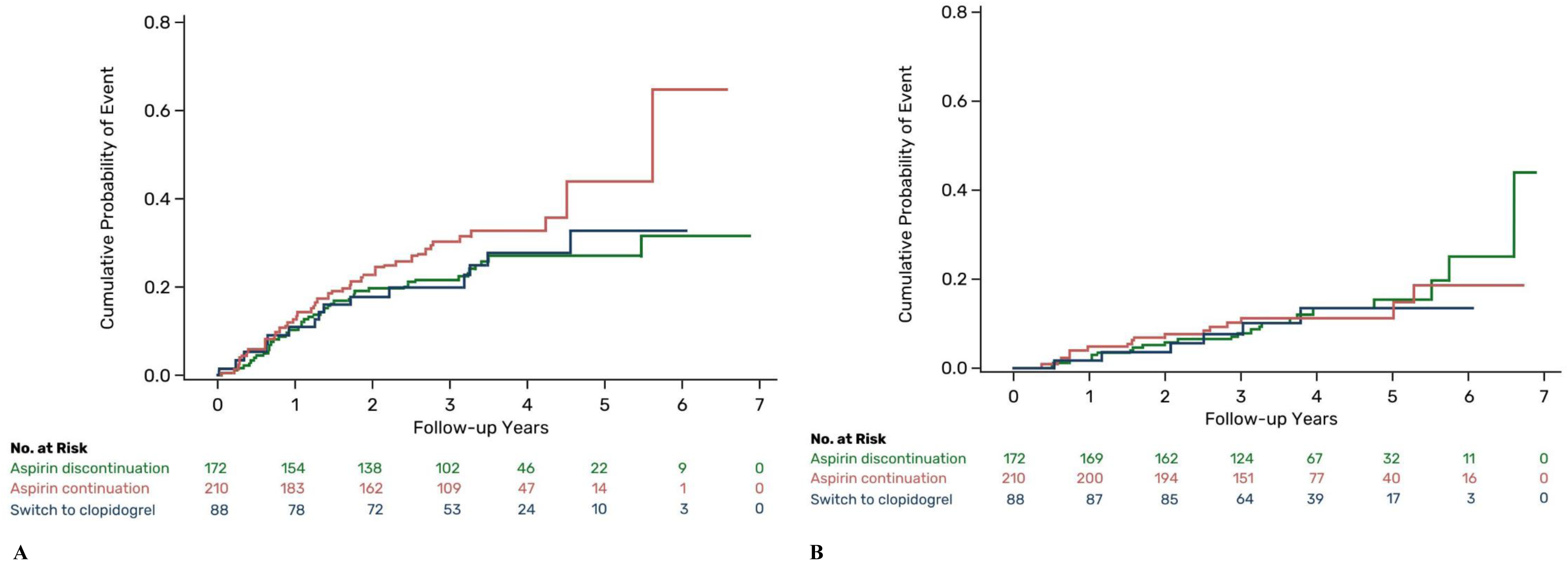
Association of subsequent antiplatelet strategies with MACEs and hospitalizations for UGIB in patients previously on aspirin for secondary CVD prevention. Kaplan–Meier failure plots show cumulative risks of major adverse cardiovascular events (A) and hospitalizations for upper gastrointestinal bleeding (B) of the three groups after inverse probability treatment weighting. Both the log-rank tests were not significant with *P* = .170. CVD, cardiovascular disease; MACE, major adverse cardiac event; UGIB, upper gastrointestinal bleeding.

**Table 1.**
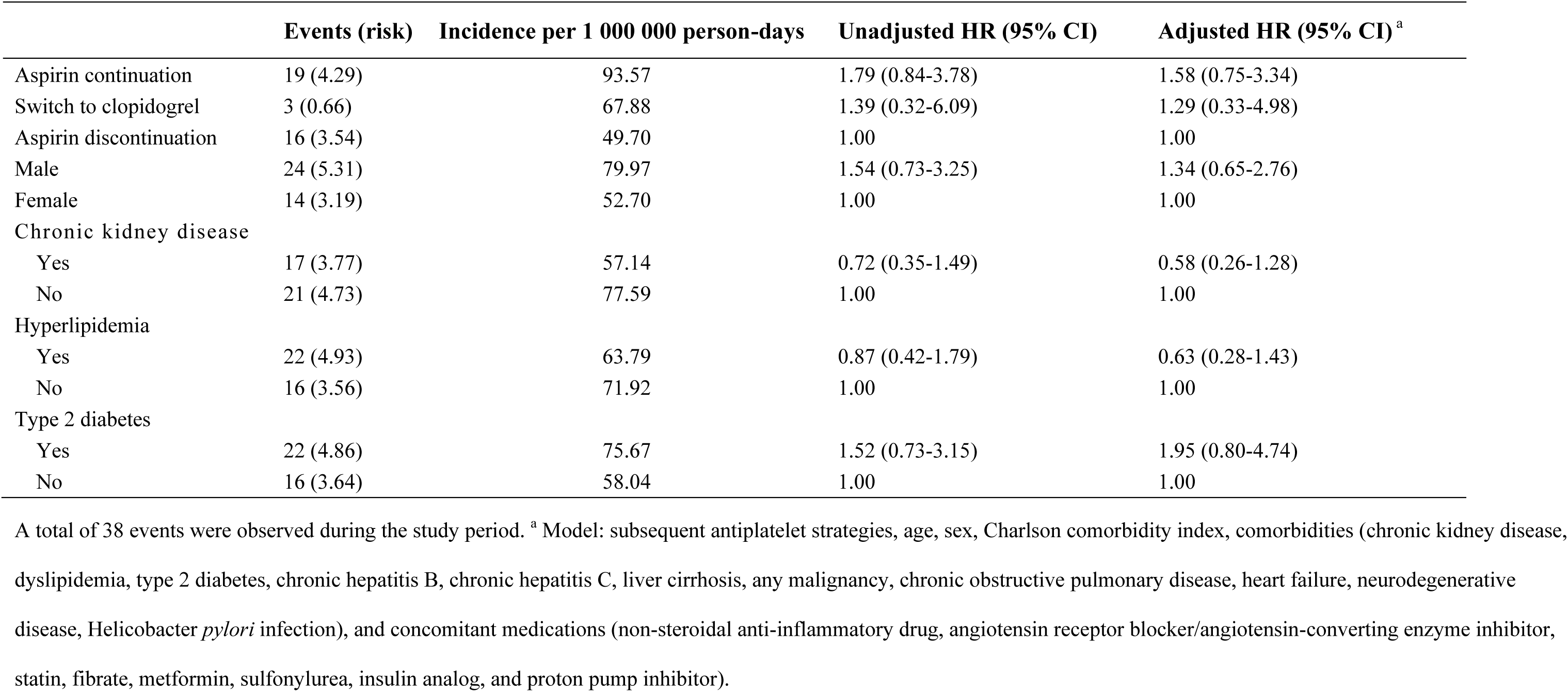
Subsequent antiplatelet strategies and incident major adverse cardiac events in patients previously on aspirin for primary cardiovascular disease prevention.

|  | Events (risk) | Incidence per 1 000 000 person-days | Unadjusted HR (95% CI) | Adjusted HR (95% CI)<sup>a</sup> |
| --- | --- | --- | --- | --- |
| Aspirin continuation | 19 (4.29) | 93.57 | 1.79 (0.84-3.78) | 1.58 (0.75-3.34) |
| Switch to clopidogrel | 3 (0.66) | 67.88 | 1.39 (0.32-6.09) | 1.29 (0.33-4.98) |
| Aspirin discontinuation | 16 (3.54) | 49.70 | 1.00 | 1.00 |
| Male | 24 (5.31) | 79.97 | 1.54 (0.73-3.25) | 1.34 (0.65-2.76) |
| Female | 14 (3.19) | 52.70 | 1.00 | 1.00 |
| Chronic kidney disease |  |  |  |  |
| Yes | 17 (3.77) | 57.14 | 0.72 (0.35-1.49) | 0.58 (0.26-1.28) |
| No | 21 (4.73) | 77.59 | 1.00 | 1.00 |
| Hyperlipidemia |  |  |  |  |
| Yes | 22 (4.93) | 63.79 | 0.87 (0.42-1.79) | 0.63 (0.28-1.43) |
| No | 16 (3.56) | 71.92 | 1.00 | 1.00 |
| Type 2 diabetes |  |  |  |  |
| Yes | 22 (4.86) | 75.67 | 1.52 (0.73-3.15) | 1.95 (0.80-4.74) |
| No | 16 (3.64) | 58.04 | 1.00 | 1.00 |

A total of 38 events were observed during the study period. <sup>a</sup> Model: subsequent antiplatelet strategies, age, sex, Charlson comorbidity index, comorbidities (chronic kidney disease, dyslipidemia, type 2 diabetes, chronic hepatitis B, chronic hepatitis C, liver cirrhosis, any malignancy, chronic obstructive pulmonary disease, heart failure, neurodegenerative disease, *Helicobacter pylori* infection), and concomitant medications (non-steroidal anti-inflammatory drug, angiotensin receptor blocker/angiotensin-converting enzyme inhibitor, statin, fibrate, metformin, sulfonylurea, insulin analog, and proton pump inhibitor).

**Table 2.** Subsequent antiplatelet strategies and incident major adverse cardiac events in patients previously on aspirin for secondary cardiovascular disease prevention.

|  | Events (risk) | Incidence per 1 000 000 person-days | Unadjusted HR (95% CI) | Adjusted HR (95% CI)<sup>a</sup> |
| --- | --- | --- | --- | --- |
| Aspirin continuation | 73 (15.49) | 322.81 | 1.47 (1.01-2.14) * | 1.58 (1.04-2.38) * |
| Switch to clopidogrel | 23 (4.96) | 228.98 | 1.05 (0.63-1.74) | 1.09 (0.63-1.87) |
| Aspirin discontinuation | 44 (9.36) | 216.53 | 1.00 | 1.00 |
| Male | 59 (12.47) | 279.13 | 1.10 (0.78-1.54) | 1.00 (0.69-1.44) |
| Female | 81 (17.34) | 254.27 | 1.00 | 1.00 |
| Chronic kidney disease |  |  |  |  |
| Yes | 78 (16.54) | 277.27 | 1.10 (0.78-1.53) | 1.01 (0.69-1.47) |
| No | 62 (13.27) | 249.36 | 1.00 | 1.00 |
| Hyperlipidemia |  |  |  |  |
| Yes | 95 (20.28) | 250.56 | 0.82 (0.57-1.17) | 0.78 (0.50-1.22) |
| No | 45 (9.53) | 298.45 | 1.00 | 1.00 |
| Type 2 diabetes |  |  |  |  |
| Yes | 90 (19.09) | 288.37 | 1.23 (0.87-1.74) | 0.87 (0.53-1.43) |
| No | 50 (10.72) | 229.70 | 1.00 | 1.00 |

A total of 140 events were observed during the study period. \* $P < .05$ . <sup>a</sup> Model: subsequent antiplatelet strategies, age, sex, Charlson comorbidity index, comorbidities (chronic kidney disease, dyslipidemia, type 2 diabetes, chronic hepatitis B, chronic hepatitis C, liver cirrhosis, any malignancy, chronic obstructive pulmonary disease, heart failure, neurodegenerative disease, *Helicobacter pylori* infection), and concomitant medications (non-steroidal anti-inflammatory drug, angiotensin receptor blocker/angiotensin-converting enzyme inhibitor, statin, fibrate, metformin, sulfonylurea, insulin analog, and proton pump inhibitor).

In the AC group who previously took aspirin for secondary CVD prevention, a higher daily dose of aspirin, compared with a lower daily dose, was associated with an increased unadjusted risk of incident MACEs (HR, 1.95; 95% CI, 1.01-3.76) (**Table S3**). When compared with AD, AC with a daily dose of aspirin ≥ the third quadrant further increased the risk of incident MACEs in secondary prevention patients (adjusted HR, 2.22; 95% CI, 1.13-4.36) (*P* for trend = .017) (**Table S4**). Sensitivity tests excluding MACEs within the first 3 months in patients previously on aspirin for primary and secondary CVD prevention also yielded similar findings (**Table S5, Table S6**), even though the positive association between AC and risk of incident MACEs in secondary prevention patients was attenuated (adjusted HR, 1.49; 95% CI, 0.99-2.28) (**Table S6**).

### Incident Hospitalizations for UGIB

During a median follow-up period of 43.4 months, 102 incident hospitalizations for UGIB occurred in the IPTW population. Treatment with a PPI was very common after a diagnosis of PUD in patients previously on aspirin for primary and secondary CVD prevention (**Table S7**). Subsequent antiplatelet strategies of patients previously on aspirin for primary CVD prevention were not associated with cumulative risks of incident hospitalizations for UGIB in Kaplan–Meier failure plots whether before IPTW (*P* = .596) (**Figure S2B**) or after IPTW (*P* = .630) (**Figure 1B**). In the multivariate Cox proportional hazards analysis of the IPTW population, neither AC (adjusted HR, 0.73; 95% CI, 0.36-1.48) nor SC (adjusted HR, 0.81; 95% CI, 0.23-2.89) was associated with risk of incident hospitalizations for UGIB in primary prevention patients, compared with AD (**Table 3**). Subsequent antiplatelet strategies in patients previously on aspirin for secondary CVD prevention were not related to the cumulative risk of incident hospitalizations for UGIB in Kaplan–Meier failure plots whether before IPTW (*P* = .953) (**Figure S3B**) or after IPTW (*P* = .170) (**Figure 2B**). Neither AC (adjusted HR, 1.14; 95% CI, 0.61-2.10) nor SC (adjusted HR, 1.07; 95% CI, 0.48-2.37) affected risks of incident hospitalizations for UGIB in secondary prevention patients, compared with AD in the multivariate Cox proportional hazards analysis of the IPTW population (**Table 4**). However, the comorbid CKD in patients previously prescribed aspirin for secondary CVD prevention increased the risk of incident hospitalizations for UGIB (adjusted HR, 2.41; 95% CI, 1.30-4.47) (**Table 4**).

**Table 3.** Subsequent antiplatelet strategies and incident hospitalizations for upper gastrointestinal bleeding in patients previously on aspirin for primary cardiovascular disease prevention.

|  | Events (risk) | Incidence per 1 000 000 person-days | Unadjusted HR (95% CI) | Adjusted HR (95% CI)<sup>a</sup> |
| --- | --- | --- | --- | --- |
| Aspirin continuation | 13 (2.79) | 59.45 | 0.68 (0.34-1.33) | 0.73 (0.36-1.48) |
| Switch to clopidogrel | 3 (0.77) | 75.46 | 0.84 (0.27-2.59) | 0.81 (0.23-2.89) |
| Aspirin discontinuation | 27 (5.97) | 85.81 | 1.00 | 1.00 |
| Male | 23 (5.20) | 76.96 | 1.04 (0.57-1.90) | 1.23 (0.63-2.42) |
| Female | 20 (4.33) | 73.22 | 1.00 | 1.00 |
| Chronic kidney disease |  |  |  |  |
| Yes | 23 (5.09) | 78.19 | 1.10 (0.60-2.00) | 0.94 (0.46-1.91) |
| No | 20 (4.44) | 72.08 | 1.00 | 1.00 |
| Hyperlipidemia |  |  |  |  |
| Yes | 24 (5.39) | 69.28 | 0.80 (0.43-1.46) | 0.70 (0.35-1.42) |
| No | 19 (4.13) | 84.69 | 1.00 | 1.00 |
| Type 2 diabetes |  |  |  |  |
| Yes | 27 (6.05) | 95.61 | 1.74 (0.93-3.24) | 2.17 (0.95-4.98) |
| No | 16 (3.47) | 54.82 | 1.00 | 1.00 |

A total of 43 events were observed during the study period. <sup>a</sup> Model: subsequent antiplatelet strategies, age, sex, Charlson comorbidity index, comorbidities (chronic kidney disease, dyslipidemia, type 2 diabetes, chronic hepatitis B, chronic hepatitis C, liver cirrhosis, any malignancy, chronic obstructive pulmonary disease, heart failure, neurodegenerative disease, *Helicobacter pylori* infection), and concomitant medications (non-steroidal anti-inflammatory drug, angiotensin receptor blocker/angiotensin-converting enzyme inhibitor, statin, fibrate, metformin, sulfonylurea, insulin analog, and proton pump inhibitor).

**Table 4.** Subsequent antiplatelet strategies and incident hospitalizations for upper gastrointestinal bleeding in patients previously on aspirin for secondary cardiovascular disease prevention.

|  | Events (risk) | Incidence per 1 000 000 person-days | Unadjusted HR (95% CI) | Adjusted HR (95% CI)<sup>a</sup> |
| --- | --- | --- | --- | --- |
| Aspirin continuation | 26 (5.45) | 89.56 | 0.95 (0.54-1.68) | 1.14 (0.61-2.10) |
| Switch to clopidogrel | 10 (2.14) | 82.74 | 0.90 (0.43-1.90) | 1.07 (0.48-2.37) |
| Aspirin discontinuation | 23 (4.89) | 96.30 | 1.00 | 1.00 |
| Male | 22 (4.68) | 85.29 | 0.91 (0.54,1.56) | 0.79 (0.43-1.45) |
| Female | 37 (7.80) | 94.40 | 1.00 | 1.00 |
| Chronic kidney disease |  |  |  |  |
| Yes | 39 (8.26) | 114.88 | 1.96 (1.12-3.42) * | 2.41(1.30-4.47) ** |
| No | 20 (4.22) | 64.32 | 1.00 | 1.00 |
| Hyperlipidemia |  |  |  |  |
| Yes | 44 (9.36) | 96.42 | 1.29 (0.71-2.35) | 1.87 (0.95-3.71) |
| No | 15 (3.12) | 77.18 | 1.00 | 1.00 |
| Type 2 diabetes |  |  |  |  |
| Yes | 33 (7.07) | 84.45 | 0.84 (0.50-1.40) | 0.72 (0.34-1.51) |
| No | 26 (5.41) | 100.59 | 1.00 | 1.00 |

A total of 59 events were observed during the study period. \* $P < .05$ ; \*\* $P < .01$ . <sup>a</sup> Model: subsequent antiplatelet strategies, age, sex, Charlson comorbidity index, comorbidities (chronic kidney disease, dyslipidemia, type 2 diabetes, chronic hepatitis B, chronic hepatitis C, liver cirrhosis, any malignancy, chronic obstructive pulmonary disease, heart failure, neurodegenerative disease, *Helicobacter pylori* infection), and concomitant medications (non-steroidal anti-inflammatory drug, angiotensin receptor blocker/angiotensin-converting enzyme inhibitor, statin, fibrate, metformin, sulfonylurea, insulin analog, and proton pump inhibitor).

## Discussion

Faced with an aging population, atherosclerotic CVD prevention is a global public health priority. Aspirin has been used for decades as a primary and secondary prevention strategy for CVD in older adults. However, accumulating evidence from trials and meta-analyses involving a large proportion of hypertensive participants has suggested that the benefits of aspirin use for primary prevention in older adults may be outweighed by the potential major bleeding risk.^7,21^ On the other hand, aspirin use for secondary CVD prevention in older hypertensive adults appears to be beneficial.^22^ Despite current recommendations for aspirin use in older patients, clinicians still struggle with decisions when aspirin users aged 65 or older develop PUD. No clinical trials have investigated the appropriateness of continuation or discontinuation of antiplatelet agents in older hypertensive adults with panendoscopy-proven PUD. The present multicenter cohort study analyzed older hypertensive adults with documented PUD using aspirin for primary and secondary CVD prevention, respectively, to investigate the outcomes of subsequent antiplatelet strategies (AD, AC, or SC) in terms of incident MACEs and hospitalizations for UGIB. The results showed that neither AC nor SC was associated with a reduced risk of incident MACEs in older hypertensive patients with panendoscopy-proven PUD previously on aspirin for primary CVD prevention, compared with AD. While SC was not related to a lowered risk of incident MACEs, AC increased the risk of incident MACEs in secondary prevention patients, compared with AD. Neither AC nor SC was associated with risk of incident hospitalizations for UGIB in primary prevention patients, compared with AD. Subsequent antiplatelet strategies were not shown to affect the risk of incident hospitalizations for UGIB in patients previously prescribed aspirin for secondary CVD prevention. Notably, the comorbid CKD in patients previously prescribed aspirin for secondary CVD prevention increased the risk of incident hospitalizations for UGIB.

Maintaining aspirin use for primary CVD prevention in older hypertensive patients with panendoscopy-proven PUD was not shown to reduce the risk of incident MACEs in our study, consistent with results from large trials studying older people with mild to moderate cardiovascular risks on aspirin for primary CVD prevention.^5–7^ The American College of Cardiology and American Heart Association Task Force guideline 2019 on the primary prevention of CVD only suggests low-dose aspirin (75-100 mg daily) administration to patients aged 40-70 years with high CVD risk and low risk of gastrointestinal bleeding.^23^ The benefit of CVD prevention associated with aspirin was also not demonstrated in the post hoc analysis of a randomized clinical trial comparing intensive and standard guideline-directed antihypertensive strategies in a primary prevention cohort of hypertensive patients at increased risk of atherosclerotic CVD but without diabetes, CVD, or CKD.^24,25^ Treating hypertension with antihypertensive medications and comprehensive prevention strategies may be more paramount than prescribing aspirin when primary CVD prevention is considered. Our results support the discontinuation of antiplatelet therapy once older hypertensive adults on aspirin for primary CVD prevention get documented PUD (**Figure 1**, **Table 1**).

On the other hand, a Swedish nationwide population-based cohort study of patients on aspirin for primary or secondary CVD prevention showed that discontinuation of aspirin in the absence of major surgery or bleeding was associated with an over 30% increased CVD risk.^26^ A small-sized randomized clinical trial of patients on low-dose aspirin for secondary CVD prevention with UGIB revealed that continuous aspirin therapy reduced the short-term mortality rate, but the study end points did not include MACEs.^17^ In the present multicenter cohort study, continuous aspirin therapy increased the risk of incident MACEs in older hypertensive adults with documented PUD using aspirin for secondary CVD prevention even after multiple adjustments (**Table 2**). In addition, a third quadrant or greater defined daily dose of aspirin in patients who continued aspirin was associated with a much higher HR for incident MACEs than AD in patients taking aspirin for secondary CVD prevention (**Table S4**). Although the positive association between AC and risk of incident MACEs in our older hypertensive patients previously on aspirin for secondary CVD prevention was attenuated in a sensitivity test excluding MACEs within the first 3 months (**Table S6**), it warrants conducting more long-term studies with information on aspirin doses actually taken.

This study also revealed that subsequent antiplatelet strategies were not associated with increased risk of incident hospitalizations for UGIB in high-risk older hypertensive adults with documented PUD previously on aspirin, whether for primary or secondary CVD prevention. In addition, PPI administration after a diagnosis of PUD is reimbursed by the National Health Insurance in Taiwan, and was therefore common in our data (**Table S7**). Although continuous PPI therapy was not observed to reduce UGIB events in an aspirin prevention trial of community-dwelling older adults,^27^ the PPI-driven protective effects against UGIB might attenuate the potential bleeding risks caused by antiplatelet drugs in older aspirin users.^28^ On the other hand, CKD is known to increase gastrointestinal rebleeding rates and higher mortality in patients with gastrointestinal bleeding.^29^ Our findings consistently showed that comorbid CKD increased the risk of incident hospitalizations for UGIB in patients previously prescribed aspirin for secondary CVD prevention, independent of PPI use (**Table 4**). To alleviate the risk of incident hospitalizations for UGIB, CKD evaluation probably counts more than antiplatelet strategies in patients previously prescribed aspirin for secondary CVD prevention. For a lower hospitalization risk for UGIB, it is never too late to curtail CKD development in patients previously on aspirin for secondary CVD prevention. Taken together, our results indicate that in the era of universal PPI use after a diagnosis of PUD, the appropriateness of continuing antiplatelet drugs should not be evaluated merely relying on UGIB risk. Instead, the actual effectiveness of continuing antiplatelet drugs for CVD prevention should be critically judged first.

Some limitations of this study should to be addressed. First, it is limited in generalizability to aspirin users other than our study population. Second, the NTUH-Integrative Medical Database does not require timely updates on residual confounding factors, including drug adherence, body mass index, smoking habits, alcohol consumption, and out-of-pocket medication history. Nevertheless, these factors are less likely to affect the clinical decisions of physicians to prescribe subsequent antiplatelet agents for the study population. To compensate for the retrospective design, we applied IPTW to balance baseline group characteristics by a weight of propensity scores.^20^ However, the baseline PPI use before panendoscopy-proven PUD in patients on aspirin for secondary CVD prevention was not balanced even after IPTW (**Table S2**). The diagnosis of PUD in patients with existing PPI treatment may cause physicians to discontinue antiplatelet therapy. Despite these concerns, this is the first multicenter cohort study examining the outcomes of antiplatelet therapy continuation after a diagnosis of PUD in older hypertensive patients previously on aspirin for CVD prevention.

## Conclusions

This pioneer multicenter study demonstrated that antiplatelet continuation did not reduce the occurrence of incident MACEs in older hypertensive patients with panendoscopy-proven PUD previously on aspirin for primary or secondary CVD prevention. Notably, continuous aspirin use after a diagnosis of PUD increased the risk of incident MACEs in secondary prevention patients, perhaps in a dose-dependent manner. In the era of universal PPI use after a diagnosis of PUD, continuation of antiplatelet agents may not increase incident hospitalizations for UGIB in primary or secondary prevention patients. It is crucial to prevent CKD development to reduce incident hospitalizations for UGIB in secondary prevention patients. Although aspirin is still recommended for secondary CVD prevention in older adults with confirmed atherosclerotic CVD,^30^ more clinical trials are warranted to justify if the benefits of continuing aspirin therapy after a diagnosis of PUD in secondary prevention patients outweigh the risks.

## Data Availability

Data not available due to ethical and legal restrictions of the NTUH-Integrative Medical Database.

## Acknowledgments

The authors appreciate statistical support from Dr. Shu-Lin Chuang, PhD, Integrative Medical Database Center, Department of Medical Research, National Taiwan University Hospital. The authors are grateful to MedCom Asia, Inc for the editing assistance of this manuscript. The authors convey our gratitude to Dr. Yi-Chen Lee, MD, Department of Community and Family Medicine, National Taiwan University Bei-Hu Branch for her collaboration during this study.

## Supplemental Material

Supplemental Methods

Figure S1–S3

Tables S1–S7

